# Building a resilient antibiotic market An econometric modelling approach to estimating revenues for a novel, broad-spectrum intravenous antibiotic

**DOI:** 10.64898/2026.03.13.26348309

**Authors:** Leela Maitreyi, Sreenath Rajagopal, Anand Anandkumar, Santanu Datta

## Abstract

India faces a mounting health crisis from antibiotic resistance, coupled with global pharmaceutical hesitancy to invest in novel antibiotic research and development (R&D), driven by complex scientific and financial hurdles. India carries one of the world’s largest absolute burdens of drug-resistant infections. The combination of a huge infectious-disease caseload, rapid urbanisation, and gaps in sanitation and primary care means that, when resistance emerges, it affects far more patients and generates a much larger pool of patients needing advanced antibiotics than in many high-income countries. Against this backdrop, demand for truly novel, broad-spectrum antibiotics in India is surging, fueled by rising multidrug-resistant infections, overstretched hospitals, and an antibiotic resistance market projected to grow rapidly over the next decade. Most countries respond with incentives and subscription models, for India, the answer lies in bold, innovative revenue strategies and in prioritising the domestic launch of novel antibiotics. This paper presents an econometric analysis of estimated valuation for a novel broad-spectrum antibiotic in India that, as a single therapeutic agent, can address several major hospital-acquired infections, including complicated urinary tract infections (cUTI), hospital-acquired pneumonia (HAP), and ventilator-associated pneumonia (VAP). The model focuses on a hypothetical “ideal” broad-spectrum intravenous antibiotic, and recommends that India pioneer market entry, highlighting financial models that maximise early revenues while still hardwiring stewardship. Launching new antibiotics first in India can catalyse robust real-world use, strengthen domestic pharma, and demonstrate that the economics of antibiotic innovation are viable. This decisive shift can transform India from a passive recipient of ageing drugs into the crucible where the next generation of life-saving antibiotics is forged, anchoring antibiotic research at the core of the country’s health security and economic resilience.

## Introduction

Launching a novel broad-spectrum antibiotic first in India is a bold and unconventional market entry. It gently challenges the usual Western-first playbook. Since the Industrial Revolution, technology and solutions have often emerged from places under the most pressure. Mass car production and the assembly line, developed in the United States, enabled people to travel across vast distances. The steam engine expanded to move goods across global trade networks. Computers and automation evolved to ease the bottlenecks caused by labour shortages in industry. India now faces some of the world’s highest levels of multidrug resistance. It may increasingly contribute to antibiotic innovation.[1–4] Necessity once again encourages invention. It allows a company to step onto the world stage within the antimicrobial resistance (AMR) challenge. Here, the stakes are high, and both impact and sustainable revenues can develop together.

### Where the fire burns the hottest

India is widely recognised as one of the major centres of AMR. The country has a large number of deaths associated with multidrug-resistant infections each year. Hospitals face pathogens that increasingly resist last-line drugs [1–5]. In this context, a genuinely novel broad-spectrum antibiotic represents more than an incremental improvement. It can influence mortality trends, intensive care unit (ICU) utilisation, and hospital economics in measurable ways. Introducing such a therapy where the disease burden is highest shapes a narrative of response in which the need is most pressing.

### From an overlooked market to a power base

For many years, India and other low- and middle-income countries (LMICs) have often entered the commercial sequence only after Western launches matured. Reversing this sequence could gradually position India as a strategic base rather than simply a secondary destination. With over a million multidrug-resistant infections annually and a resistance-focused market already valued in the billions and growing, India may support a meaningful, premium-positioned antibiotic franchise rather than merely tail-end market [6, 7]. Being among the early innovators in this space allows companies to help shape treatment guidelines, stewardship practices, and payer expectations, rather than compete in a crowded, highly genericised market.

### A live laboratory for proof and prestige

An India-first launch means the drug is evaluated under demanding real-world conditions: busy ICUs, patients presenting late in disease progression, polymicrobial infections, and high levels of resistance. Demonstrating clinical value in this environment can generate a strong body of real-world evidence that later supports Western clinical trial dossiers, payer discussions, and pricing considerations [8–12]. Success in such settings can also bring reputational benefits, positioning the company not only as an innovator but as one willing to demonstrate its therapy in challenging healthcare environments.

### Economics that bend in your favour

India’s development, manufacturing, and commercialisation costs are typically lower than those in the US or Europe. This can strengthen the economic case for innovation [13–16]. Incremental clinical success in high-income countries can translate into meaningful system-level benefits in India. These benefits include shorter hospital stays, fewer salvage regimens, and reduced catastrophic out-of-pocket spending. Such outcomes support differentiated pricing and maintain an image of responsible, patient-centred innovation.

### Writing a new global script

Choosing India as an early launch market may signal a different type of antibiotic company. Such a company collaborates closely with global health organisations, AMR initiatives, and access-oriented partnerships while maintaining a sustainable commercial outlook [17–19]. If implemented carefully, with stewardship embedded, thoughtful pricing structures, and regional expansion, India could become a launchpad. This would broaden expectations of where innovation emerges. Leadership in antibiotic development can flow from Bengaluru and Mumbai to Boston and Berlin, instead of only the other way around.

In a country like India, where the AMR burden is enormous and hospital systems are already strained, feeding local epidemiology, utilisation, and cost data into econometric models can directly translate the AMR burden into projected revenues, budget impact, and ROI for a novel antibiotic launch [1, 3, 4, 8, 11, 20–22]. These approaches expose the fallacy behind the belief that “there is no market for new antibiotics.” Instead, they show that the problem is not a lack of demand, but a lack of clear, quantified business cases in the highest-burden countries (Fig.1). A predictive econometric model is indispensable for estimating revenue in the antibiotics industry. It elevates forecasting from heuristic extrapolation to a transparent, mechanistic representation of how infections, resistance, and clinical decision-making create demand [21–26].

**Fig. 1:**
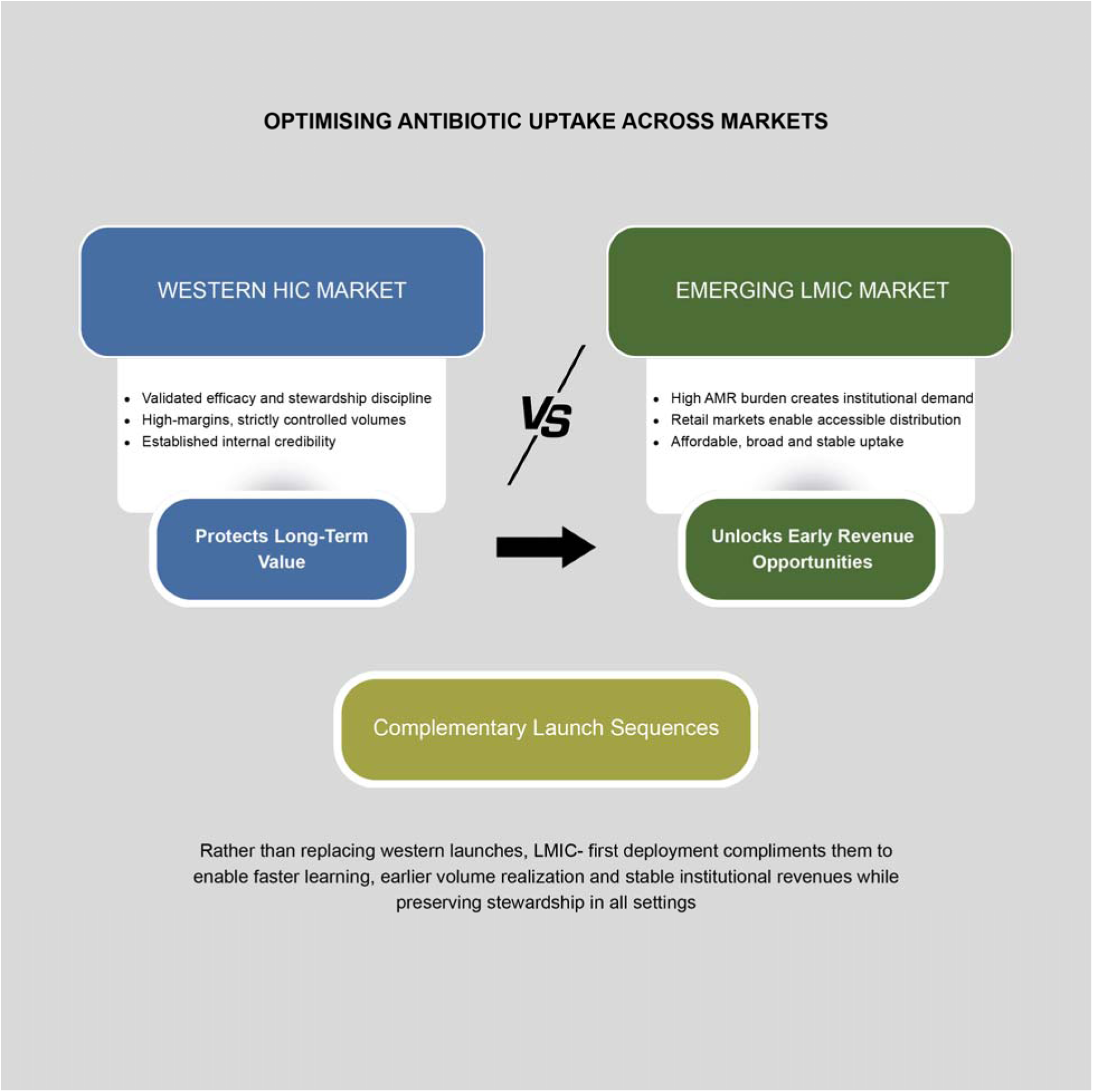
Antibiotic Uptake Across Markets.

For a soon-to-be-launched novel antibiotic, such a model can assimilate country-level incidence of multi-drug resistant (MDR)/extensively drug-resistant (XDR) pathogens, current standard-of-care pathways, stewardship constraints, and pricing assumptions to generate internally coherent projections of volume and revenue over time. This structure also enables rigorous scenario analysis, varying guideline inclusion, formulary tiering, competitive entries, and access policies, to delineate a credible range of commercial outcomes rather than a single, opaque point estimate.

For a preclinical, hypothetical “ideal” broad-spectrum intravenous (IV) antibiotic positioned in an India-first context, this approach is even more valuable. The model can ingest local surveillance data, ICU and ward utilisation, length-of-stay distributions, and cost structures. It converts a qualitative AMR burden into a quantified future addressable market, budget impact, and risk-adjusted return on investment. The model helps dismantle the narrative that “there is no market for new antibiotics.” It reveals that the real deficit is the absence of robust business cases anchored by multiple models in high-burden settings [1, 20–24, 27].

## Methods

The paper focuses on building predictive revenue models for the launch of the novel broad-spectrum antibiotic Drug A and a hypothetical “ideal” broad-spectrum IV antibiotic in India. These models were built around real clinical burden indicators, including AMR prevalence, ICU admission rates, infection incidence, and hospital-acquired infection frequencies, to estimate structurally necessary treatment volumes [1, 4, 21, 25, 28, 29]. The key indications represented mathematically in these models are complicated urinary tract infection (cUTI), hospital-acquired pneumonia (HAP) and ventilator-acquired pneumonia (VAP). Long-term AMR trend projections are modelled for both approved antibiotics and the candidate therapy to assess revenue durability and demand stability under increasing resistance pressures. The combined framework produces revenue estimates grounded in mandatory clinical use, guideline inclusion, and patient access and affordability, delivering realistic and financeable forecasts. Because drug revenues are driven by biological uncertainty, policy behaviour, and market dynamics simultaneously, and no single model can capture all three, the paper uses multi-model forecasting (Fig.2): Epidemiological models estimate the number of patients who need the drug based on disease burden and resistance patterns. This model projects the addressable patient pool for novel broad-spectrum antibiotics by integrating India-specific epidemiology of cUTI and HAP/VAP [1, 21, 23, 25, 30–32]. The input data are shown in the supplementary materials.

**Fig. 2:**
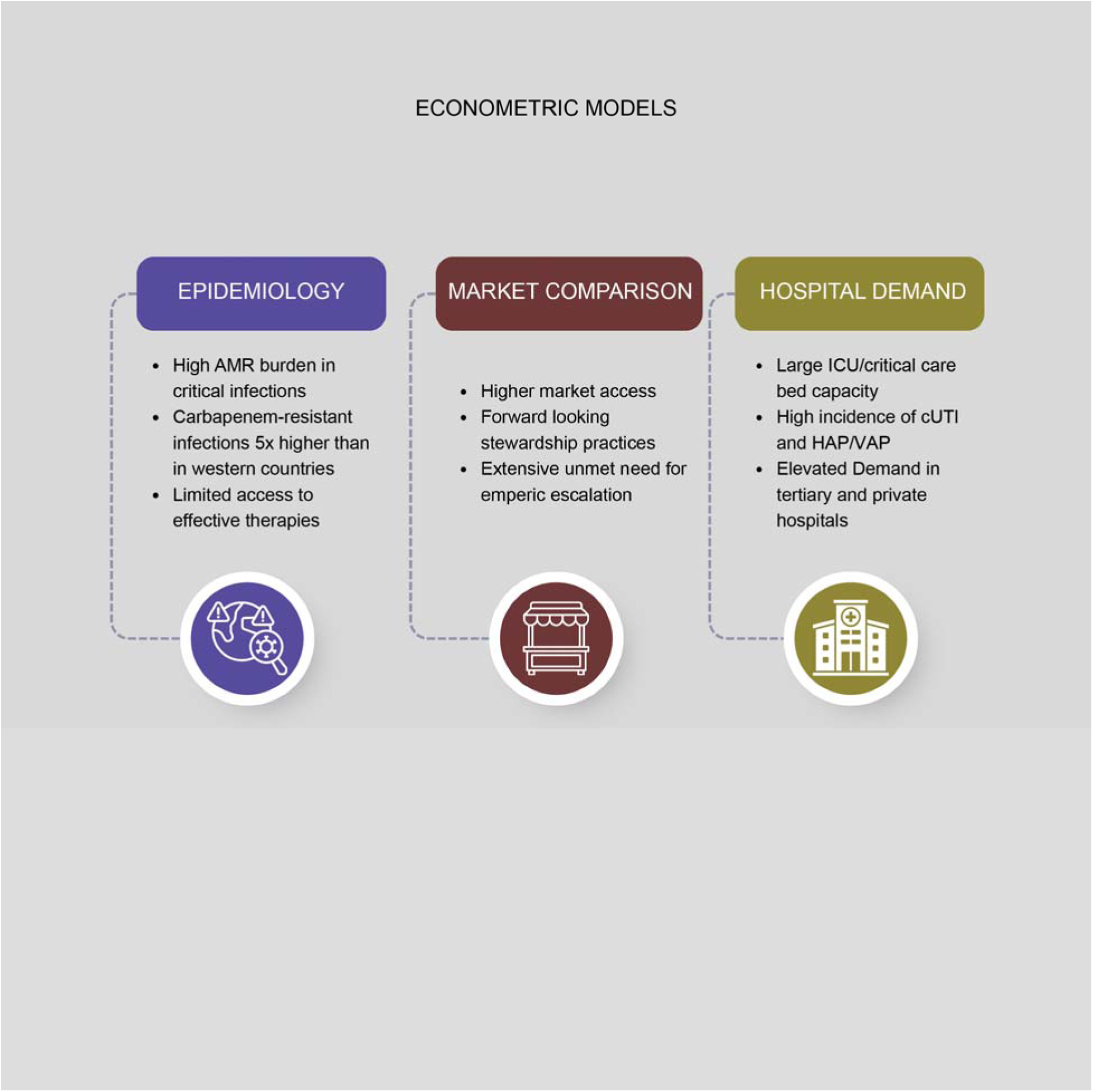
Econometric models for revenue estimations in India.

A market-based model was built to estimate revenue for new broad-spectrum antibiotics using a market segmentation approach that divides India’s total antibiotics market by IV formulation, novelty (innovator vs generic), and AWaRe category (Fig.3) (Access/Watch/Reserve). IV antibiotics dominate hospital settings (e.g., 50-60% value in tertiary care for severe MDR cases), innovators command premiums in Reserve (1% volume but high value due to scarcity), versus generics flooding Access/Watch (Watch: 34% volume)[16, 33–35]. The input data are shown in the supplementary materials.

**Fig. 3:**
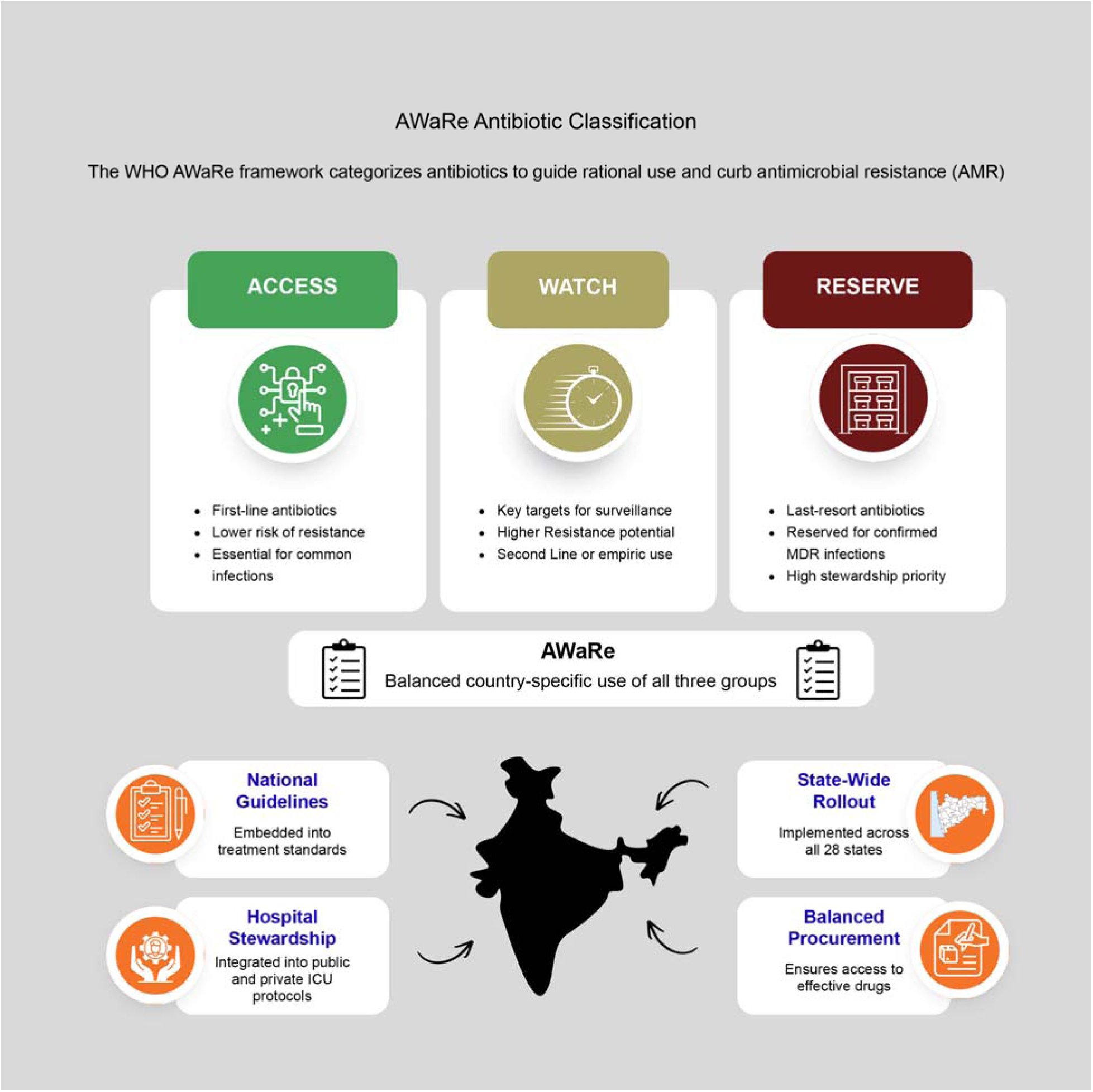
AWaRe Antibiotic Classification.

Hospital Bed Model estimates antibiotic revenue by anchoring demand to physical healthcare capacity rather than to total patient counts alone. The model accounts for the total number of functional hospital beds (public and private), segmented into general and ICU beds. Bed occupancy rates were applied to estimate active patient-days per year. Standard-of-care (SOC) guideline eligibility filters were applied to identify cases in which the new antibiotic would be clinically indicated. Treatment volume was computed by multiplying the number of eligible cases by the duration of the new antibiotic course. Beds are fixed infrastructure assets, budgets are allocated per bed, and infections scale with bed-days , so revenue forecasts become capacity-linked, budget-anchored, and stewardship-safe, rather than inflated by retail prescription assumptions. This model turns antibiotic revenue into infrastructure-linked cash flow. The input data are shown in the supplementary materials. [1, 17, 20, 21, 28, 29, 36–38].

### Input data and model structure

Within each of the 3 models, 3 scenarios were built in: Base case (Pragmatic, highly conservative), upside case (Best case, still conservative), downside case (Cynical) Indian pharmaceutical multi-year financial metrics and performance indicators were curated from open-source reports for 10 companies. From this data, the key financial metrics were calculated as %: Cost of goods (COGS), Selling, general & administrative expenses (SG&A), R&D, Capital expenses (CAPEX). The mean and median financial metrics for these companies were used as model parameters. All model assumptions, formulae & quantitative input data are provided in the supplementary.

### Two case studies were modelled in this paper

#### Drug A-With the following specifications

Spectrum: Novel beta-lactamase/beta-lactamase inhibitor (BL/BLI) class antibiotic designed and developed to treat life-threatening, multidrug-resistant (MDR) Gram-negative bacterial infections, which are a major global health challenge. The mechanism allows this drug to effectively combat "superbugs," including those resistant to carbapenems (the current gold standard treatments)

Positioning: For high-risk cUTI and HAP/VAP in ICU settings Launch Timeline: To launch in India by 2026

*Hypothetical “Ideal” broad-spectrum IV antibiotic was considered to fulfil the following specifications:*

Spectrum: Developed to address multidrug-resistant (MDR) Gram-negative bacterial infections, all CRE, Carbapenem-resistant *Acinetobacter baumannii* (CRAB), Carbapenem-resistant *Pseudomonas aeruginosa* (CRPA) and is a novel bacterial topoisomerase inhibitor (NBTI) class

Positioning: For high-risk cUTI and HAP/VAP in ICU settings

Estimated Launch Timeline: To launch in India by 2031

The models for these two antibiotics were built using the discounted cash flow (DCF)methodology. DCF [39–42]models for new antibiotics forecast value by discounting future cash flows from projected sales at a high-risk Weighted Average Cost of Capital (WACC) 10-15%), capturing India-specific dynamics such as market penetration, stewardship caps, and MDR caseload elasticity [1, 17, 21, 39–47].

Core Structure: 10-15 year explicit forecasts project revenue ramp using a $100M & $50M sunk cost for market entry & preclinical assets resp. (sales/marketing heavy upfront), capex (initial launch infrastructure), and corporate tax. Terminal value assumes 3-5% perpetual growth amid sustained AMR.

Key Parameters: Peak sales (India ICU focus), SoC failure substitution (meropenem/Ceftazidime / Avibactam)[1, 17, 47–49], sensitivity to penetration, pricing elasticity.

Using multiple model types allowed us to triangulate revenue from clinical necessity, pricing sensitivity, adoption speed, and resistance evolution simultaneously, producing forecasts that are resilient to biological, regulatory, and economic uncertainty — something a single-model forecast cannot deliver.

## Results

All simulated scenarios are presented in the Supplementary Materials.

The results (Fig.4)(Fig.5) show the model’s outputs across different simulated scenarios. 27 scenarios are in the mean category & median category each

**Fig. 4:**
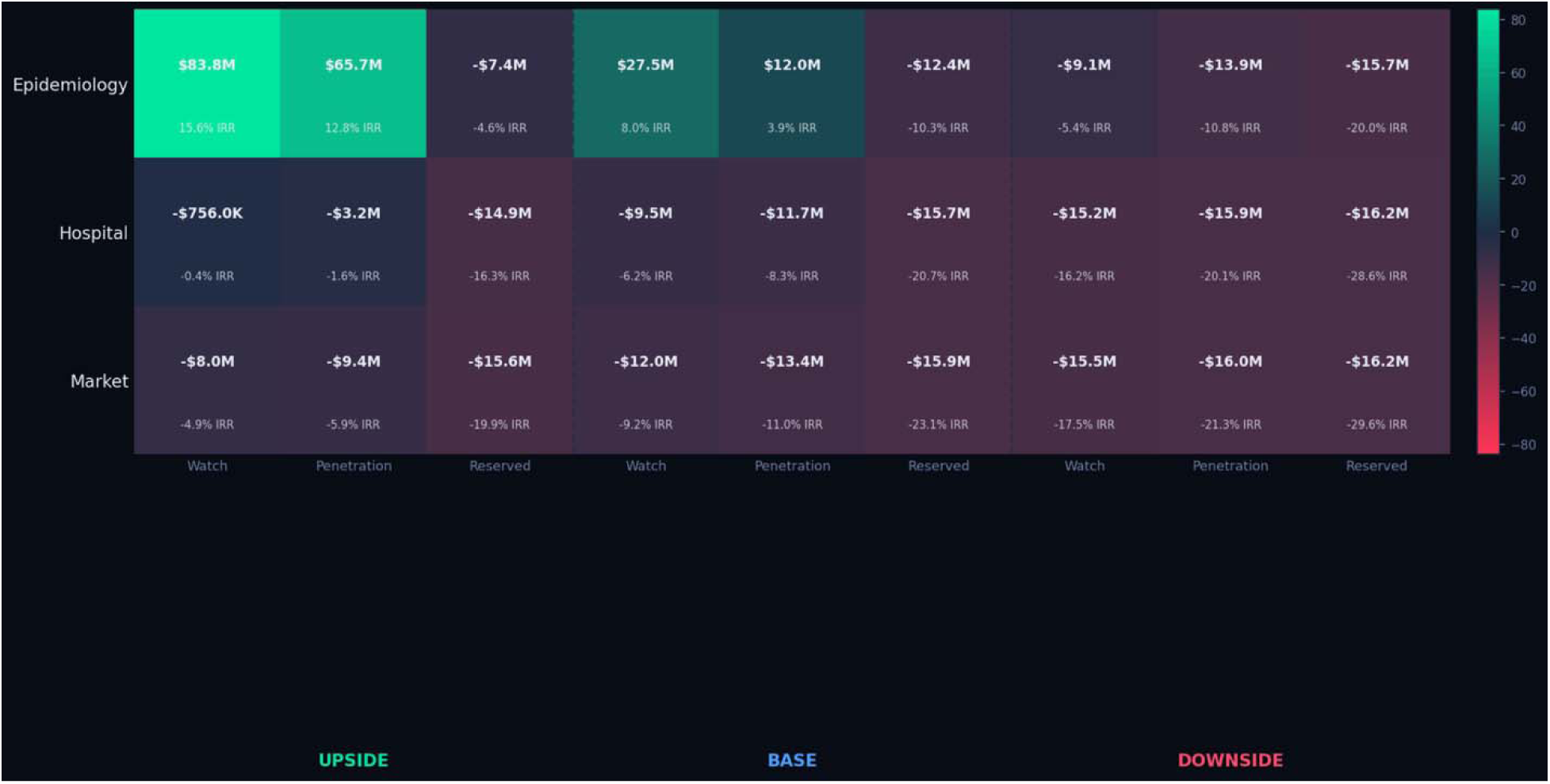
Valuation & IRR estimations for a broad-spectrum preclinical asset across different model scenarios.

**Fig. 5:**
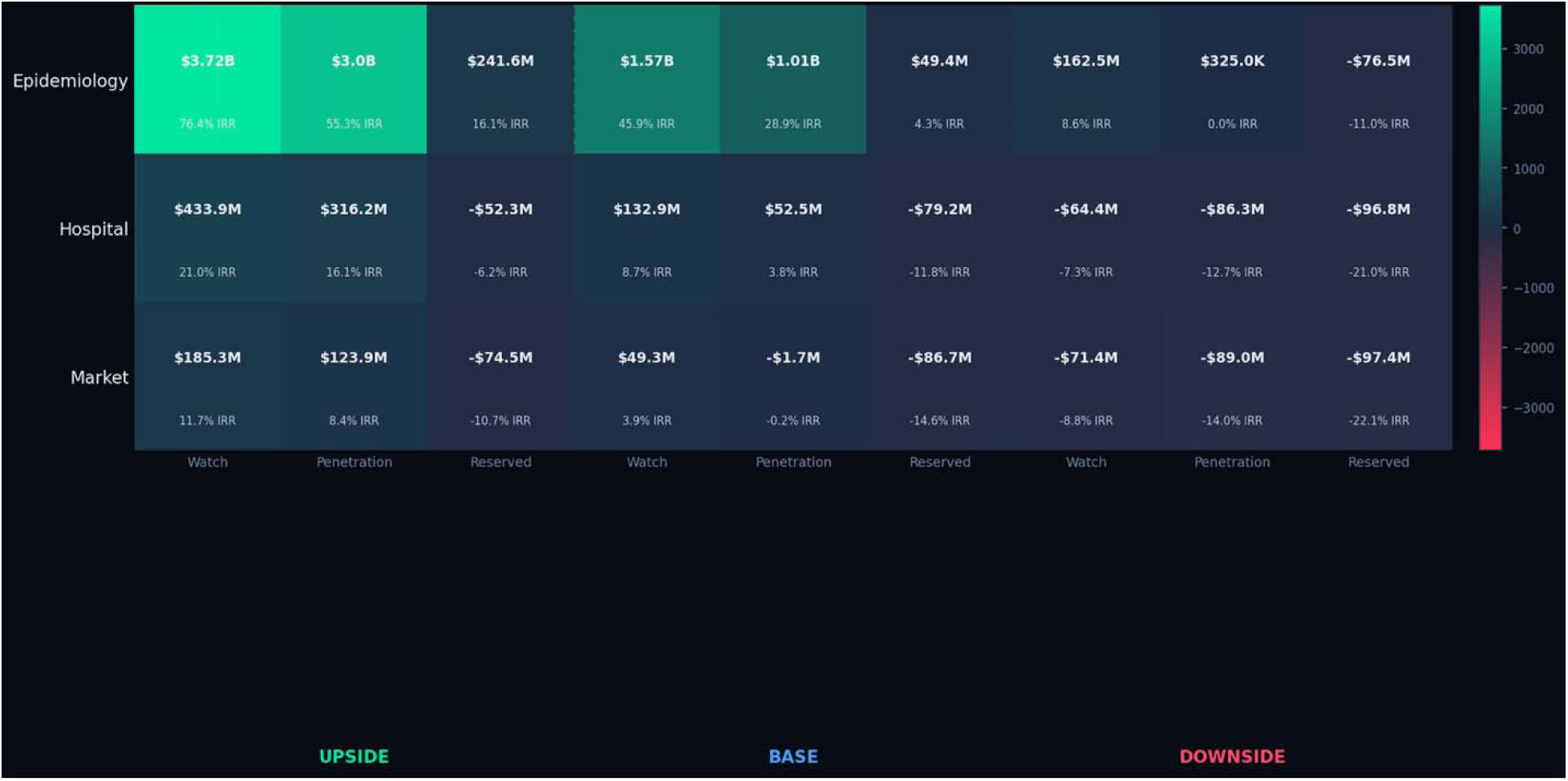
Valuation & IRR estimations for market-ready Drug-A across different model scenarios.

Best of the median and mean scenarios are illustrated in the heat maps (Fig. 4, Fig.5) The same modelling framework was used for both case studies.

This allows the outcomes to be compared under a consistent analytical approach.

Model evaluation under the specified scenarios reveals distinct patterns in system behaviour, summarised as follows:

Case-1: Preclinical hypothetical “ideal” broad-spectrum IV antibiotic 54 scenarios were simulated. Scenarios are combinations of base/pragmatic, best and worst cases across the 3 models explained in the methods section. Out of the 54 scenarios highlighted, peak revenue is in the range of $13.8 million to $1406 million. 8 scenarios show net positive NPV & positive ROI, measured through IRR in the range of 1.57%-15.65%

Case-2: Soon-to-be-launched Drug A

54 scenarios were simulated Out of the 54 scenarios highlighted, peak revenue is in the range of $14 million to $1434 million, 28 scenarios show net positive NPV & positive ROI, measured through IRR in the range of 0.02%-76%

Key takeaways from the 2 case studies

For the preclinical asset: IRR is mostly negative because an antibiotic company has all the costs but none of the monetizable value yet, while facing significant time, failure, and pricing risks.In financial terms, its projected cash flows are highly negative and heavily discounted, long capital lock-up before any revenue, extreme probability-of-failure multipliers, stewardship caps revenue because the drug is intentionally positioned to be underused and cannot be aggressively marketed. Resistance shrinks product life, so terminal revenue is structurally smaller than oncology, rare disease, or chronic drugs. Additionally, our models are shown to be extremely conservative in terms of market penetration rate-very conservative at 8%,10% &12 % of SOC displacement rates, COGS/CAPEX-highly aggressive estimate, using the financial metrics of the 10 Indian pharma companies that have small molecules and biologics in their pipeline. The actual COGS for a small-molecule drug is far lower and is expected to be even lower for a non-BL/BLI class. Very conservative per-course treatment cost at $1000 shown. Very aggressive development cost shown at $50Million from R&D to market

For a soon-to-be-launched asset: IRR is positive because once an antibiotic asset reaches the clinical stage, the entire financial structure of the project changes, because biological uncertainty becomes smaller with time. Clinical-stage assets have already cleared the most stringent scientific filters. As the probability of success rises, expected terminal cash flows jump dramatically, instantly lifting the IRR. Time to Revenue Shrinks. Time to revenue is considerably shorter for clinical-stage assets. While early-stage discovery programs may take 12–15 years to reach commercialisation, clinical-stage assets are typically only 3–6 years away from potential revenue generation. Accordingly, our models apply a 10% discount rate for market-ready assets and a 15% discount rate for preclinical assets, ensuring conservative and risk-adjusted DCF estimates. Every clinical milestone doubles or triples the enterprise value.

Manufacturing Economics are Visible. COGS, scale curves, and pricing bands become known, making margins financeable. This makes revenue models credible and lowers discount rates.

Clinical-stage assets reach cash breakeven much faster, dramatically improving IRR even if lifetime revenues are modest. As a result, the lens moves from “Long, uncertain, distant upside” to “Shorter, de-risked, credible cashflows” and IRR becomes positive even in this paper. This paper aims to address factors that make a compelling case for developing, trialling, and launching a novel broad-spectrum antibiotic in India first, while achieving reasonable revenues and protecting stewardship.

## Discussion

Preclinical antibiotic assets typically demonstrate scenarios, and negative IRRs because they face long development timelines, high biological failure risk, and distant, heavily discounted future cash flows. Their key bottlenecks include scientific uncertainty, lack of contractable demand, limited manufacturing visibility, and structurally capped revenue potential due to stewardship-driven volume suppression [22, 43–45, 50–53]. In contrast, clinical-stage antibiotic companies demonstrate rapidly improving NPVs and positive IRRs, driven by reduced development risk, shorter time-to-market, eligibility for early procurement contracts, and clearer manufacturing economics [22, 24, 44, 45, 50, 52, 54]. Crucially, India represents a particularly effective launch platform for a novel antibiotic, enabling both meaningful public health impact and commercial sustainability. High levels of antimicrobial resistance create urgent clinical demand, while the country’s manufacturing cost advantages allow affordable pricing without compromising margins.[1–5, 35]. Together, these structural features transform uncertain retail demand into predictable institutional cashflows, lowering discount rates and accelerating payback periods—thereby converting previously negative-NPV antibiotic assets into financeable, positive-IRR infrastructure-grade businesses.

Our probabilistic forecasting framework integrates AMR prevalence, ICU and hospital bed capacity, stewardship-driven prescribing constraints, cost sensitivity, and market-penetration dynamics to simulate multiple real-world launch scenarios for a novel broad-spectrum IV antibiotic in India. By explicitly modelling uptake against high-resistance standard-of-care (SOC) pathways and protocol-restricted prescribing, the framework estimates mandatory clinical demand rather than aspirational market size. Domestic manufacturing cost curves are layered to define affordable yet commercially sustainable pricing corridors. Resistance trend projections and guideline-integration timelines are incorporated to test the durability of long-term demand and the stability of revenue. We further apply multi-model sensitivity analysis, stress-testing worst-case, best-case, and pragmatic scenarios across market archetypes, financial metrics (NPV, IRR, payback), penetration rates, cost structures, and forecasting methodologies. Across all modelled cases, the analysis consistently demonstrates predictable institutional volumes, budget-compatible margins, and accelerated payback periods, providing a robust, data-driven basis for positioning India as an optimal global launchpad for this product.

Our probabilistic forecasting models highlight the following:

WATCH-category antibiotics generate the highest revenues because they are used broadly as empiric therapy during epidemic and outbreak conditions, when first-line ACCESS options fail due to resistance, creating rapid, large-scale, hospital-wide uptake that drives exceptional but episodic revenue surges. This is the most ambitious scenario and an unlikely model to be adopted under normal infection conditions in India. This revenue behaviour is unique to epidemic dynamics, in which the WATCH drugs sit at the exact inflexion point between ACCESS failure and RESERVE restriction, allowing high-volume deployment before stewardship ceilings are triggered.

RESERVE-category antibiotics, by contrast, show structurally lower revenues because they are tightly restricted to salvage therapy, limiting volumes even in high-burden AMR environments despite their higher per-course prices.

Market penetration assumptions of 6%, 10%, and 12% remain conservative because existing SOC carbapenems exhibit high and rising resistance across Indian ICU pathogens, forcing clinicians to escalate empiric therapy rapidly — structurally expanding the eligible treatment population. In high-resistance hospital settings, empiric escalation often exceeds these penetration bands, particularly during ventilator-associated pneumonia clusters, underscoring the modelled penetration levels’ intentional caution.

A total development-to-launch cost of approximately USD 50 million is a reasonable estimate for a non-BL/BLI antibiotic, reflecting shorter clinical pathways, narrower chemistry complexity, LMIC-based trial efficiencies, and domestic manufacturing scale that materially compresses development and commercialisation expenditure.

India’s scale dwarfs perception: The absolute burden translates into substantial revenue potential even at modest pricing, if positioning is empiric for ICU.

Quantifies “no market” myth: Models show demand exists, but requires multi-model business cases

### Why launch in india first?

India provides a distinctive setting in which stewardship, access, and commercial sustainability for novel antibiotics can coexist. The country’s high burden of antimicrobial resistance and hospital-acquired infections creates durable baseline demand for clinically appropriate use, even under responsible stewardship. At the same time, India’s cost-efficient pharmaceutical manufacturing ecosystem enables innovators to achieve sustainable margins through scale rather than high per-course pricing. Early inclusion in national treatment guidelines can further secure durable first-mover advantages through institutional procurement and formulary adoption.

Together, these factors position India not only as a viable early launch market but also as a platform for generating real-world evidence that may inform adoption in other low- and middle-income settings and potentially de-risk entry into high-income markets.

This study evaluates the principal economic drivers of such a launch strategy. Specifically, it estimates the burden of hospital-acquired infections in India that could be addressed by a novel broad-spectrum intravenous antibiotic and quantifies the addressable patient cohort after applying stewardship criteria. The analysis models stewardship caps that preserve long-term drug durability while maintaining commercial viability and assesses pricing architecture and revenue trajectories over a fifteen-year horizon. Using conservative, base, and optimistic scenarios, we estimate peak revenues, break-even points, and internal rates of return, while incorporating cost-of-goods assumptions to guide sustainable per-course pricing.

Several factors lie beyond the scope of the present analysis. The model does not incorporate policy incentives or financing mechanisms such as grants, tax credits, market-entry rewards, or subscription models that could further strengthen antibiotic development economics. Intellectual property and data-exclusivity frameworks, post-approval stewardship commitments, and potential revenues from out-licensing or royalty agreements are also excluded. In addition, the analysis does not model expansion of this approach to other low- and middle-income markets or the potential for Indian market validation to influence launches in Europe or North America.

Finally, the study focuses on intravenous formulations and does not estimate revenue potential for oral, inhalable, or topical antibiotic products.

## Conclusions

While much of the AMR economics literature portrays antibiotic markets as structurally broken, India offers a setting where stewardship, access, and commercial sustainability can coexist. Its high burden of hospital-acquired infections and drug-resistant Gram-negative pathogens creates a durable baseline demand, even under responsible stewardship, making it a viable early-launch environment for novel intravenous broad-spectrum antibiotics [1–4, 22, 38].

In many Indian tertiary hospitals, antibiotic prescribing is guided by clinical protocols and supervision, supporting appropriate treatment volumes within stewardship frameworks.

Stewardship, therefore, stabilises institutional demand rather than eliminating revenue. This effect is reinforced by India’s expanding publicly financed healthcare infrastructure: procurement systems and insurance schemes such as Ayushman Bharat convert fragmented patient spending into pooled, budget-anchored demand, lowering revenue volatility while maintaining affordability through negotiated pricing [1, 17, 24, 36, 37, 43, 47, 55–59].

India’s manufacturing ecosystem further aligns access with innovation, enabling the production of novel antibiotics at low marginal cost while remaining commercially viable through scale.

Early inclusion in national treatment guidelines can also create first-mover advantages through physician standardisation and institutional procurement preference, positioning India as both a revenue base and a validation platform within a broader launch sequence [15, 38, 60, 61].

Taken together, India suggests that antibiotic markets need not remain structurally broken. Where stewardship, pooled procurement, manufacturing scale, and guideline integration converge, access and stewardship can coexist with predictable revenues. Rising incomes and an estimated 100–300 million upper-middle-class consumers further support uptake of novel antibiotics priced above generics, challenging perceptions of India as a uniformly low-ability-to-pay market [62–66].

## Data Availability

All data produced in the present work are contained in the manuscript

## Acknowledgement

We thank the leadership team at Bugworks Research, including Dr. Balasubramanian V [COO], Dr. Shahul Hameed [CSO], Dr. Vasan Sambandamurthy [SVP], and Dr.Vasanthi Ramachandran [VP] for their encouragement and institutional support during the development of this work.

## Notes

### Competing Interest Statement

The authors have declared no competing interest.

### Funding Statement

This study did not receive any funding

### Summary of Updates

The title has been revised to Building a resilient antibiotic market An econometric modelling approach to estimating revenues for a novel, broad-spectrum intravenous antibiotic

