## supplementary for "Building a resilient antibiotic market An econometric modelling approach to estimating revenues for a novel, broad-spectrum intravenous antibiotic"

The Supporting Information describes the model assumptions, parameter values, and the full set of scenarios simulated in this analysis.

### Key assumptions for the models

#### CASE 1: Preclinical Hypothetical “Ideal” Broad-spectrum IV Antibiotic

- Model start year: 2020
- Launch year (India): 2031
- Analysis horizon: 25 years
- Corporate tax: 0.25%
- Discount rate (WACC): 15%
- Indications modelled: cUTI, HAP, VAP
- Within each of the 3 models, 3 scenarios were built in:
  - **Base case (Pragmatic, highly conservative)**
  - **Upside case (Ambitious)**
  - **Downside case (Cynical)**
- Population end-point methods used (Model types)
  - **Epidemiology**
  - **Market diffusion/penetration**
  - **Hospital admission/ICU beds**
- **Indications used for the modelling are:**
  - **cUTI**
  - **HAP**
  - **VAP**

##### Inputs:

- **Base case cost/course/patient:** $1000
- **Base case market penetration to displace existing SOC:** 10%
- **Market share at the peak penetration is considered to be** 10%
- **Access/payer discount is captured at** 20%
- **Total cost of development, trial & launch only in India:** $50Million
  - The probability of success shown is from one phase to another
  - Peak revenue estimation is shown between 4-6 years
  - COGS is shown as 38% of the sales
  - SG&A is shown as 33% of the sales
  - R&D is shown as 6% of the sales
  - CAPEX is 6% of the sales
  - These metrics were calculated from 10 Indian pharma companies financials, using open-source reports
- **Total antibiotic market estimate shown is for India alone**
  - This is further divided by product type IV Vs Oral, generic Vs branded, human Vs veterinary
  - Further estimations classified into Watch and Reserve category used from the AWaRe classification of Antibiotics use in India
- **Hospital model**
  - Shows all govt and private hospital beds
  - Total beds for the computation is shown as 50% of the actual total, as a conservative measure
  - ICU beds are reported to be 5% of the total beds
  - Number of ICU beds occupied in a given day as shown as 80%
  - Ventilator usage shown in 30% of the bed occupied by the patients
  - VAP cases are reported to be 6.4 patients for every 1000
  - Non-VAP cases are reported to be 60% of HAP cases
- **Epidemiology model**
  - Average incidence of cUTI is computed at 30% of the total patient pool
  - 85% of the patients are reported to have sought medical intervention
  - Of those, only 50% sought hospitalization
  - Of those, only 60% sought Antibiotic treatment
  - Based on the Kerala formularies and registries, 1% of the total Antibiotics fall under the “Reserve” category
  - Based on the Kerala formularies and registries, 11% of the total Antibiotics fall under the “Watch” category
  - Ventilator days/year is 530
- Valuation is computed using the DCF model
- Mean and median values were used for the financial metrics from these companies and used as toggles for each of the models
- WACC(discount rate) is used at 15% (highly aggressive)
- Overall probability of success is shown at 16.99%

#### CASE 2: Soon-to-be-launched DRUG A

- Model start year: 2025
- Launch year (India): 2026
- Analysis horizon: 18 years
- Corporate tax: 0.25%
- Discount rate (WACC): 10% because it is a clinical stage asset
- Indications modelled: cUTI, HAP, VAP
- Within each of the 3 models, 3 scenarios were built in:
  - **Base case (Pragmatic, highly conservative)**
  - **Upside case (Ambitious)**
  - **Downside case (Cynical)**
- Population end-point methods used (Model types)
  - **Epidemiology**
  - **Market diffusion/penetration**
  - **Hospital admission/ICU beds**
- **Indications used for the modelling are:**
  - **cUTI**
  - **HAP**
  - **VAP**

##### Inputs:

- **Base case cost/course/patient:** $1000
- **Base case market penetration to displace existing SOC:** 10%
- **Market share at the peak penetration is considered to be** 10%
- **Access/payer discount is captured at** 20%
- **Total cost of development, trial & launch globally:** $100Million
  - A comparable Indian Pharma company data showed $500 million spend from preclinical to market
  - The pipeline for this spend had 6 assets
  - Per asset cost computed ~=$84 Million (In India, US and China)
  - Two models for sunk cost can be computed:
    - Empirical model 1:We assume $100Million as a very conservative number (Model used)
    - Empirical model 2: $50Million as a more realistic ballpark for India alone(Can be Simulated)
  - The probability of success shown is from one phase to another
  - Peak revenue estimation is shown between 4-6 years
  - COGS is shown as 38% of the sales
  - SG&A is shown as 33% of the sales
  - R&D is shown as 6% of the sales
  - CAPEX is 6% of the sales
- These metrics were calculated from 10 Indian pharma companies financials, using open-source reports
- **Total antibiotic market estimate shown is for India alone**
  - This is further divided by product type IV Vs Oral, generic Vs branded, human Vs veterinary
  - Further estimations classified into Watch and Reserve category used from the AWaRe classification of Antibiotics use in India
- **Hospital model**
  - Shows all govt and private hospital beds
  - Total beds for the computation is shown as 50% of the actual total, as a conservative measure
  - ICU beds are reported to be 5% of the total beds
  - Number of ICU beds occupied in a given day as shown as 80%
  - Ventilator usage shown in 30% of the bed occupied by the patients
  - VAP cases are reported to be 6.4 patients for every 1000
  - Non-VAP cases are reported to be 60% of HAP cases
- **Epidemiology model**
  - Average incidence of cUTI is computed at 30% of the total patient pool
  - 85% of the patients are reported to have sought medical intervention
  - Of those, only 50% sought hospitalization
  - Of those, only 60% sought Antibiotic treatment
  - Based on the Kerala formularies and registries, 1% of the total Antibiotics fall under the “Reserve” category
  - Based on the Kerala formularies and registries, 11% of the total Antibiotics fall under the “Watch” category
  - Ventilator days/year is 530
- Valuation is computed using the DCF model
- Mean and median values were used for the financial metrics from these companies and used as toggles for each of the models
- WACC(discount rate) is used at 10% (clinical asset)
- Overall probability of success is shown at 100%(clinical asset)
